# Myocardial Perfusion in Patients with Primary Mitral Regurgitation Due to Mitral Valve Prolapse

**DOI:** 10.64898/2026.10.04.26364711

**Authors:** Klara Lodin, Jannike Nickander, Anne Wang, Jacob Widaeus, Lauge Østergaard, Lars H Lund, Peter Svenarud, Magnus Dalén, Sara Tehrani, Marcus Carlsson, Bahira Shahim

## Abstract

**Background:** Mitral regurgitation (MR) is a common complication of mitral valve prolapse (MVP) and increases left ventricular (LV) preload, promotes LV remodeling and hypertrophy, but its impact on myocardial perfusion and coronary microvascular dysfunction remains unclear.

**Objectives:** To assess myocardial perfusion using, for the first time, quantitative cardiac magnetic resonance (CMR) perfusion mapping in patients with significant MR due to MVP.

**Methods:** Patients with significant primary MR, without coronary artery disease were prospectively enrolled and compared to age- and sex-matched controls without MR. All underwent 1.5T CMR including cine imaging, adenosine stress perfusion, native T1, native T2, extracellular volume (ECV) mapping and late gadolinium enhancement (LGE). Myocardial perfusion reserve (MPR) was calculated as the ratio of stress to rest myocardial perfusion.

**Results:** In total 33 patients and 33 controls were included (age 56±13 vs. 58±12 years, 46% women in both groups). Patients with MR had increased LV mass and volumes, higher native T1 and ECV and a higher prevalence of LGE (36% vs 0%). No differences were observed in global myocardial perfusion at rest (0.88±0.27 vs. 0.85±0.30, p=0.52), at stress (2.6±0.56 vs. 2.8±0.73 ml/min/g, p=0.29) or for MPR (3.1±0.75 vs. 3.4±1.0, p=0.24). In age- and sex-adjusted linear regression, higher NYHA class was associated with lower MPR (β =–0.60, p=0.04).

**Conclusion:** In patients with significant MR due to MVP, myocardial perfusion appears preserved despite LV volume overload. However, lower MPR in symptomatic patients suggests that microvascular dysfunction may emerge in a more advanced disease stage.

---

Mitral valve prolapse (MVP) is the most common cause of primary mitral regurgitation (MR), which is initially characterized by a compensated hemodynamic state but over time leads to progressive left ventricular (LV) remodelling and ultimately heart failure (1). In other conditions with chronic volume overload, elevated LV preload has been linked to impaired myocardial perfusion due to increased myocardial rest perfusion and thereby a reduced myocardial perfusion reserve (MPR), implicating coronary microvascular dysfunction (CMD) (2). However, in primary MR it remains uncertain whether myocardial perfusion is altered and, if so, whether any impairment reflects CMD per se or secondary hemodynamic effects of volume overload.

Quantitative cardiovascular magnetic resonance (CMR) adenosine stress perfusion mapping enables non-invasive assessment of myocardial perfusion and MPR, with good agreement compared with positron emission tomography (3), while multiparametric CMR provides complementary evaluation of cardiac anatomy, function, and tissue characteristics (4–6). CMR-derived MR volume and regurgitant fraction (RF) have demonstrated prognostic value in patients with primary MR and are increasingly recognized as important markers for risk stratification and timing of surgery (7). To date, myocardial perfusion in patients with MVP and significant MR has not been studied using quantitative CMR perfusion mapping.

Thus, the aim of this study was to assess global myocardial perfusion in patients with moderate or severe MR due to MVP using adenosine stress CMR quantitative perfusion mapping, and to compare these findings with healthy controls with no MR.

## Methods

### Study population

This prospective observational study included patients with MVP and with moderate or severe MR who were consecutively enrolled in the ongoing MITRA-VT study (NCT06255457) after referral for mitral valve surgery. The study investigates the molecular mechanisms underlying MVP in distinct patient subsets, and how mitral valve surgery affects CMR biomarkers and symptom burden over time, as well as clinical outcomes, including ventricular arrhythmias. Exclusion criteria were coronary artery disease (ruled out by either coronary computed tomography or coronary angiography), rheumatic valve disease, mitral stenosis, previous valve intervention, significant aortic stenosis or regurgitation, endocarditis, or known primary cardiomyopathy. Control subjects were free from MVP or other significant valvular heart disease. Patients with primary MR underwent CMR between March 2024 and

November 2025, while controls were examined between June 2016 and August 2025. Control subjects had no prior angina, myocardial infarction or revascularization. Controls were selected to match the patient cohort as closely as possible with respect to age, sex, and cardiovascular risk factors, including hypertension, hypercholesterolemia, diabetes, and smoking status. The control cohort comprised participants from three previously conducted studies. Twenty-five controls were randomly sampled from the general population as part of the Swedish Cardiopulmonary Bioimage Study (SCAPIS) (8), whereas controls from the QUACK (n=5) (5) and SMINC (n=3) (6) studies were recruited through public advertisements and participated on a voluntary basis. All procedures were granted ethical approval by the Swedish Ethical Review Authority.

### Image acquisition

Patients and controls underwent the same CMR protocol. CMR was performed supine at 1.5T (MAGNETOM Aera or Sola scanner, Siemens Healthineers, Forchheim, Germany). Hematocrit and creatinine were determined prior to CMR. Full-coverage retrospective electrocardiographic-gated balanced steady-state free precession cine imaging was acquired in short-axis and 3 long-axis slices.

Quantitative perfusion maps were acquired in 3 short-axis slices (basal, midventricular, apical) using first-pass perfusion imaging during adenosine infusion (140 µg/kg/min or increased if inadequate response of heart rate or blood pressure to 210 µg/kg/min) (Adenosine, Life Medical AB, Stockholm, Sweden), and at rest during injection of intravenous contrast (0.05 mmol/kg, gadobutrol; Gadovist, Bayer AB, Berlin, Germany) (9, 10). Adenosine response was assessed by heart rate response and symptoms. Adenosine and contrast were administered in separate cannulas.

Native T1 maps were acquired in one midventricular short-axis slice with confirmation in one four-chamber view using an electrocardiographic-gated modified look-locker inversion 5s(3s)3s recovery research sequence. Postcontrast T1 maps were acquired after a contrast bolus (0.15 mmol/kg; gadobutrol) with the same image positions as the native T1 map. Extracellular volume (ECV) maps were generated from native T1 map and postcontrast T1 map and calibrated by the hematocrit. Native T2 maps were acquired in one midventricular short-axis slice using a T2-prepared product sequence (Siemens MyoMaps product sequence). Postcontrast late gadolinium enhancement (LGE) images were acquired in short-axis and 3 long-axis slices using a free-breathing phase-sensitive inversion recovery sequence with balanced steady-state free precession single-shot readout.

### CMR image analysis

Images were anonymized and analysed offline using Segment software version 4.1.0.1 (Medviso AB) (11). LV volumes and mass were acquired by manually delineating epi- and endocardial borders of the cine short axis-stack in end diastole and end systole. LV volumes and mass were indexed to body surface area calculated with the Mosteller formula with LV mass expressed as the LV mass index (LVMI). MR volume was calculated as LV stroke volume (LVSV) − aortic forward flow volume, and RF was calculated as MR volume / LV stroke volume. MR severity was categorized according to previously proposed CMR-specific thresholds for primary MR as mild (RF<20%), moderate (RF 20– 39%), and severe (RF≥40%) (12). The LV end-diastolic volume to LVM ratio (EDV/LVM) was calculated and used as a measure of the extent of adaptive LV hypertrophy and thus a reflection of global end-diastolic wall stress (13). LGE was considered present if visually identified on two contiguous short-axis slices or confirmed in one short-axis slice and one additional orthogonal imaging plane, and its extent was quantified in millilitres and as a percentage of total LVM. Native T1, native T2, ECV and perfusion maps were analysed by manually delineating epi- and endocardial borders in respective short-axis stack. To avoid contamination from the blood pool and adjacent tissues, a 10% erosion margin was set for endo- and epicardial borders. Segmental values were acquired in a 16 segment LV model adapted from the American Heart Association (14). Global native T1, native T2, ECV, and perfusion values were acquired by averaging the segmental values. Myocardial perfusion reserve (MPR) was calculated as the ratio of stress to rest perfusion. Total coronary flow at rest and during stress was calculated by multiplying myocardial rest and stress perfusion (ml/min/g) by LV mass (g), yielding an estimate of absolute coronary blood flow (ml/min).

### Assessment of mitral valve characteristics

MVP was diagnosed according to current guidelines in the parasternal long-axis view by observing a systolic displacement of the mitral leaflet into the left atrium (LA) of at least 2 mm from the mitral annular plane (15).The MVP phenotype was subdivided as single-leaflet (anterior or posterior) or bileaflet prolapse.

Subsequently, MVP was further stratified as either Barlow’s disease or “other ethiology”, including fibroelastic deficiency (FED), FED+, and forme fruste. Mitral annular disjunction (MAD) was defined as the distance between the insertion point of the posterior mitral leaflet and the LA–LV junction, visualized during both systole and diastole to distinguish it from pseudo-MAD (16). Although CMR is the reference standard, subtle features of MAD and Barlow disease can still be challenging to assess on CMR alone. Therefore, echocardiograms were systematically reviewed alongside CMR studies to reduce the risk of missed abnormalities and to strengthen the diagnostic classification. Transthoracic or transesophageal echocardiograms were performed a median of 61.5 days (interquartile range [IQR], 46.0–87.5 days) prior to CMR.

### Statistical analysis

Continuous data were presented as mean and standard deviation and categorical data as number with percentage. Patients and controls were compared with the independent t test or Mann Whitney U test depending on normality and chi square test or Fisher exact test as appropriate for categorical data. Linear regression analyses were performed among patients with MVP to assess associations of clinical and CMR variables with myocardial rest and stress perfusion, and MPR. Each variable was assessed in separate models, first unadjusted and subsequently adjusted for age and sex. The results of the linear regression were presented as unstandardized coefficients (β), along with 95% confidence intervals (CI) and p-values. Statistical tests were two-tailed and p<0.05 was considered significant. SPSS statistical software version 31 (IBM) was used for statistical analysis.

## Results

### Clinical characteristics and hemodynamic data

A total of 66 individuals were included: 33 patients with MVP and significant MR, and 33 age- and sex-matched controls. Baseline demographics, comorbidities, body surface area, heart rate, and laboratory values were comparable between groups (all p>0.05; Table 1).

**Table 1.** Baseline characteristics in controls and patients with MR.

| Characteristics | Controls<br>n=33 | Patients with MR<br>n=33 | p-value |
| --- | --- | --- | --- |
| Age, years | 57.6±12.3 | 55.7±13.0 | 0.31 |
| Sex, female | 46% (15) | 46% (15) | 1.00 |
| BMI, kg/m<sup>2</sup> | 24.9±3.6 | 24.7±4.5 | 0.98 |
| Serum creatinine, µmol/l | 76.2±14.8 | 77.7±14.6 | 0.64 |
| Hematocrit | 0.41±0.04 | 0.42±0.06 | 0.51 |
| NTproBNP, pg/ml | N/A | 400.6±693.6 |  |
| Hypercholesterolemia | 9% (3) | 9% (3) | 1.00 |
| Hypertension | 15% (5) | 18% (6) | 0.74 |
| Diabetes | 3% (1) | 3% (1) | 1.00 |
| Smoking status |  |  | 0.60 |
| Never | 79% (26) | 79% (26) |  |
| Prior | 18% (6) | 12% (4) |  |
| Current | 3% (1) | 9% (3) |  |
| NYHA class |  |  |  |
| I | N/A | 42% (14) |  |
| II | N/A | 42% (14) |  |
| III | N/A | 15% (5) |  |
| IV | N/A | 0% (0) |  |
| Mitral valve characteristics |  |  |  |
| Barlow's disease | N/A | 48% (16) |  |
| Other etiology | N/A | 52% (17) |  |
| MAD | N/A | 6% (2) |  |
| Posterior leaflet prolapse | N/A | 61% (20) |  |
| Anterior leaflet prolapse | N/A | 12% (4) |  |
| Bileaflet prolapse | N/A | 27% (9) |  |

**Abbreviations:** BMI – body mass index, MAD – mitral annular disjunction, MR – mitral regurgitation, NYHA – New York Heart Association.

Within the MR group, the majority were in New York Heart Association (NYHA) functional class I or II (I: 42%, II: 42%, III: 15%, IV: 0%). Mitral valve pathology included Barlow’s disease (48%) and other ethiologies (52%), with bileaflet prolapse present in 27%, posterior leaflet prolapse in 61% and anterior leaflet prolapse in 12% (Table 1).

Hemodynamic parameters are presented in Table 2. Compared with controls, patients with MR exhibited higher LV mass and larger LV volumes, with significantly higher LVMI, LVEDVI, LVESVI, and LVSVI (all p<0.001). The EDV/LVM-ratio and EF were also higher in patients with MR, whereas cardiac index was decreased in patients with MR (2.7±0.50 vs. 3.4±0.64 L/min/m^2^, p<0.001) Among patients with MR, 15 (46%) had moderate MR and 18 (55%) had severe MR, with a mean mitral RF of 42.5±13.1%.

**Table 2.** CMR findings in controls and patients with MR.

| CMR findings | Controls<br>n=33 | Patients with MR<br>n=33 | p-value |
| --- | --- | --- | --- |
| BSA, m<sup>2</sup> | 1.9±0.21 | 1.9±0.23 | 0.98 |
| Heart rate rest, bpm | 66.0±10.4 | 68.2±15.0 | 0.61 |
| LVM, g | 106.2±28.6 | 129.4±37.9 | 0.01 |
| LVMI, g/m<sup>2</sup> | 54.9±10.9 | 66.6±14.0 | <0.001 |
| EDV, ml | 169.6±45.0 | 237.5±62.5 | <0.001 |
| EDVI, ml/m<sup>2</sup> | 88.2±19.4 | 123.3±25.5 | <0.001 |
| ESV, ml | 70.6±22.5 | 93.0±33.1 | 0.003 |
| ESVI, ml/m<sup>2</sup> | 36.7±10.3 | 48.2±14.9 | <0.001 |
| SV, ml | 99.0±25.0 | 144.5±39.0 | <0.001 |
| SVI, ml/m<sup>2</sup> | 51.5±10.7 | 75.1±16.9 | <0.001 |
| EF, % | 58.8±5.3 | 61.0±8.0 | 0.03 |
| Cardiac index, L/min/m<sup>2</sup> | 3.4±0.64 | 2.7±0.50 | <0.001 |
| EDV/LVM ratio | 1.7±0.25 | 2.0±0.29 | <0.001 |
| RF, % | N/A | 42.5±13.1 |  |
| MR severity, degree |  |  |  |
| Moderate | N/A | 46% (15) |  |
| Severe | N/A | 55% (18) |  |
| Native T1 global, ms | 998.6±38.6 | 1021.9±31.0 | 0.01 |
| Native T2 global, ms | 48.0±2.2 | 48.7±1.9 | 0.21 |
| ECV global, % | 24.5±1.8 | 27.3±3.5 | <0.001 |
| LGE presence | 0 (0%) | 36% (12) | <0.001 |
| LGE, ml | 0.0±0.0 | 1.0±1.9 | 0.004 |
| LGE, % | 0.0±0.0 | 0.94±1.8 | 0.004 |
| Perfusion rest, ml/min/g | 0.85±0.30 | 0.88±0.27 | 0.52 |
| Total coronary rest flow, ml/min | 88.3±31.2 | 112.8±47.9 | 0.02 |
| Perfusion stress, ml/min/g | 2.8±0.73 | 2.6±0.56 | 0.29 |
| Total coronary stress flow, ml/min | 289.5±87.0 | 336.3±118.8 | 0.11 |
| MPR | 3.4±1.0 | 3.1±0.75 | 0.24 |

**Abbreviations:** BSA – body surface area, ECV – extracellular volume, EDV – end diastolic volume, EDVI – end diastolic volume index, EF – ejection fraction, ESV – end systolic volume, ESVI – end systolic volume index, LGE – late gadolinium enhancement, LVM – left ventricular mass, LVMI – left ventricular mass index, MPR – myocardial perfusion reserve, MR – mitral regurgitation, RF – mitral regurgitant fraction, SV – stroke volume, SVI – stroke volume index.

### Myocardial perfusion

Table 2 presents global perfusion values. Myocardial rest perfusion was available in all 33 patients with MR and 30 controls, while myocardial stress perfusion was available in 28 patients with MR and 32 controls. There were no differences in myocardial rest perfusion (0.88±0.27 vs. 0.85±0.30 mL/min/g, p=0.52) or myocardial stress perfusion (2.6±0.56 vs. 2.8±0.73 mL/min/g, p=0.29) between patients with MR and controls (Figure 1). However, total coronary rest flow was significantly higher in patients with MR compared with controls (112.8±47.9 vs. 88.3±31.2 mL/min, p=0.02). A similar, numerical higher, but statistically non-significant trend was observed for total coronary stress flow (336.3±118.8 vs. 289.5±87.0 mL/min, p=0.11). There was no significant difference in MPR (3.1±0.75 vs. 3.4±1.0, p=0.24), and only two patients with MR had an MPR<2.0. Patients with MR with NYHA class > I had lower MPR compared to patients with MR with NYHA class I (2.9±0.50 vs. 3.5±0.87, p=0.01) (Figure 2).

**Figure 1.**
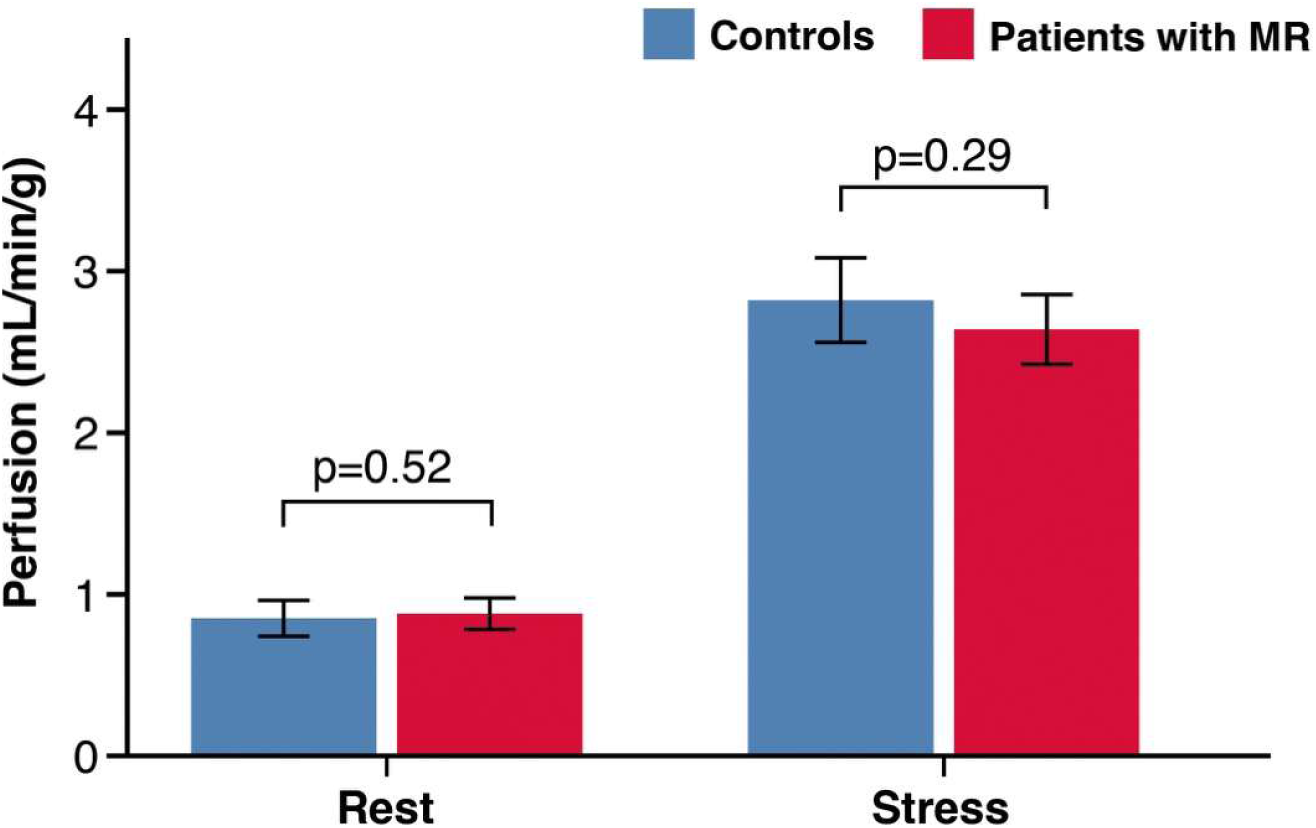
Myocardial perfusion in controls and patients with primary MR.

**Figure 2.**
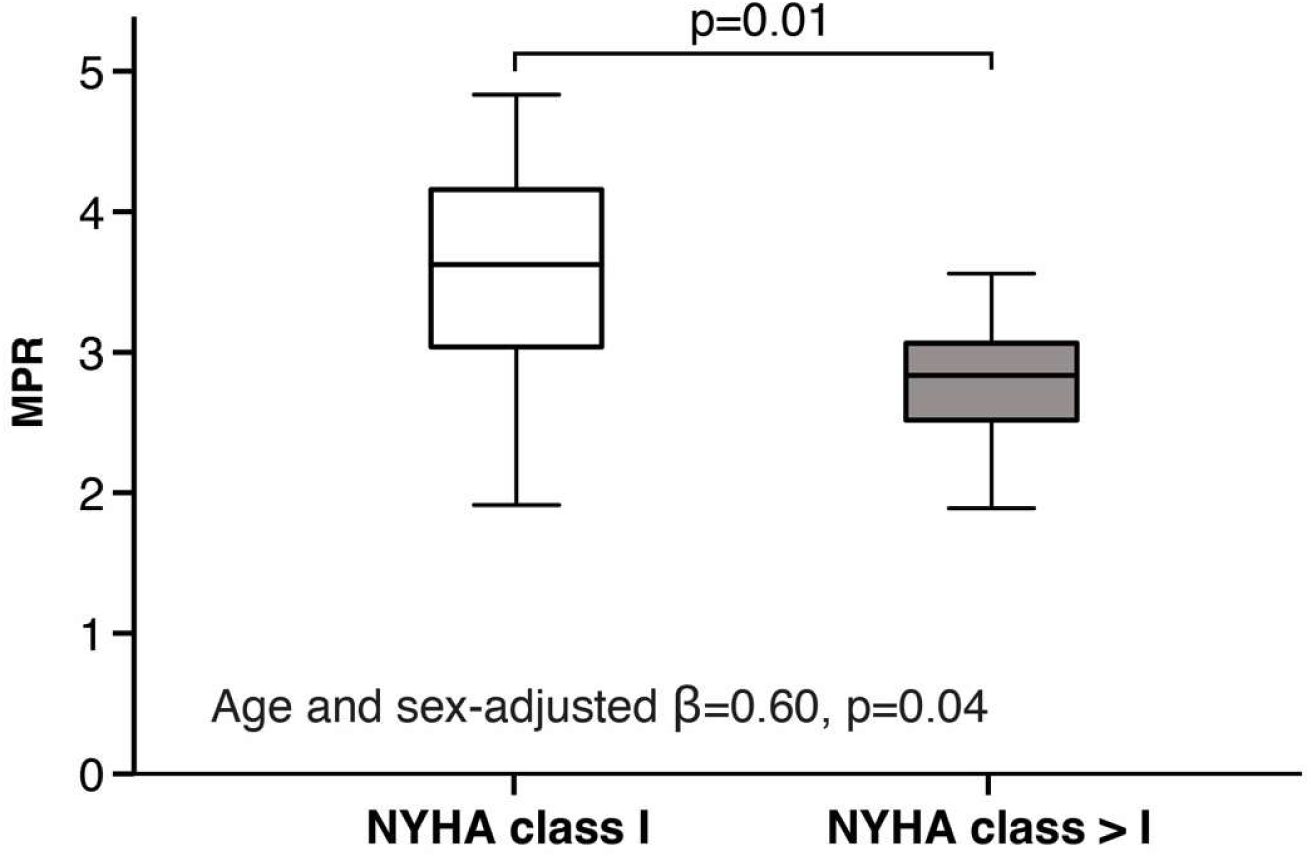
MPR and symptomatic status in patients with primary MR.

### Tissue characteristics

Table 2 presents global values of native T1, native T2, ECV, and the presence of LGE. Global native T1 values were slightly elevated in patients with MR compared with controls (1021.9±31.0 vs. 998.6±38.6 ms, p=0.01) and global ECV was also higher in patients (27.3±3.5 vs. 24.5±1.8%, p<0.001). Non-ischemic, minor LGE was present in 36% of MR patients, but absent in controls (p<0.001).

### Determinants of myocardial perfusion in primary MR

As summarized in Table 3, in crude linear regression analyses, higher age was associated with lower MPR (β=–0.03, 95% CI –0.05 to –0.002; p=0.03). Higher NYHA class was associated with lower MPR (β=–0.68, 95% CI –1.21 to –0.14; p=0.02). Higher heart rate was associated with borderline lower myocardial stress perfusion (β=–0.01, 95% CI –0.03 to 0.00; p=0.05) and lower MPR (β=– 0.02, 95% CI –0.04 to –0.001; p=0.04).

**Table 3.** Associations between clinical and CMR characteristics and myocardial perfusion in patients with MR.

| Clinical and CMR characteristics | Perfusion rest, Crude $\beta$ (95% CI). p-value | Perfusion stress, Crude $\beta$ (95% CI). p-value | MPR, Crude $\beta$ (95% CI). p-value | Perfusion rest, Adjusted $\beta^*$ (95% CI). p-value | Perfusion stress, Adjusted $\beta^*$ (95% CI). p-value | MPR, Adjusted $\beta^*$ (95% CI). p-value |
| --- | --- | --- | --- | --- | --- | --- |
| Age, years | 0.01 (−0.002, 0.01), p=0.12 | 0.001 (−0.02, 0.02), p=0.90 | −0.03 (−0.05, −0.002), p=0.03 |  |  |  |
| Sex, female | 0.10 (−0.10, 0.29), p=0.32 | 0.28 (−0.15, 0.71), p=0.19 | −0.25 (−0.84, 0.34), p=0.39 |  |  |  |
| BMI, kg/m<sup>2</sup> | −0.02 (−0.04, 0.01), p=0.19 | −0.03 (−0.08, 0.02), p=0.26 | 0.02 (−0.05, 0.09), p=0.56 | −0.01 (−0.03, 0.02), p=0.46 | −0.02 (−0.08, 0.04), p=0.46 | −0.003 (−0.07, 0.07), p=0.94 |
| NTproBNP, pg/ml | 0.00 (0.00, 0.00), p=0.81 | 0.00 (0.00, 0.00), p=0.41 | 0.00 (0.00, 0.00), p=0.70 | 0.00 (0.00, 0.00), p=0.32 | 0.00 (−0.001, 0.00), p=0.27 | 0.00 (0.00, 0.001), p=0.60 |
| Hyperlipidemia | −0.05 (−0.39, 0.29), p=0.77 | −0.60 (−1.43, 0.22), p=0.14 | −0.85 (−1.96, 0.25), p=0.12 | −0.06 (−0.42, 0.30), p=0.75 | −0.50 (−1.38, 0.38), p=0.25 | −0.90 (−1.98, 0.18), p=0.10 |
| Hypertension | −0.05 (−0.31, 0.20), p=0.68 | −0.35 (−0.96, 0.27), p=0.26 | −0.49 (−1.32, 0.34), p=0.23 | −0.13 (−0.39, 0.13), p=0.32 | −0.37 (−1.00, 0.26), p=0.24 | −0.33 (−1.15, 0.48), p=0.41 |
| Diabetes | −0.20 (−0.77, 0.38), p=0.49 | 0.05 (−1.15, 1.24), p=0.94 | 0.77 (−0.80, 2.34), p=0.33 | −0.02 (−0.63, 0.59), p=0.94 | 0.20 (−1.13, 1.53), p=0.76 | 0.07 (−1.62, 1.75), p=0.94 |
| Smoking status | 0.02 (−0.14, 0.13), p=0.81 | −0.10 (−0.44, 0.24), p=0.55 | −0.27 (−0.72, 0.17), p=0.22 | 0.01 (−0.15, 0.17), p=0.92 | −0.08 (−0.44, 0.27), p=0.63 | −0.20 (−0.64, 0.24), p=0.36 |
| NYHA class | 0.05 (−0.15, 0.25), p=0.59 | −0.27 (−0.70, 0.17), p=0.22 | −0.68 (−1.21, −0.14), p=0.02 | 0.004 (−0.20, 0.21), p=0.97 | −0.44 (−0.89, 0.02), p=0.06 | −0.60 (−1.17, −0.04), p=0.04 |
| Heart rate, bpm | −0.001 (−0.01, 0.01), p=0.88 | −0.01 (−0.03, 0.00), p=0.05 | −0.02 (−0.04, −0.001), p=0.04 | 0.001 (−0.01, 0.01), p=0.81 | −0.02 (−0.03, −0.001), p=0.03 | −0.03 (−0.04, −0.01), p=0.01 |
| LVM, g | −0.001 (−0.004, 0.002), p=0.50 | −0.002 (−0.01, 0.004), p=0.47 | 0.003 (−0.01, 0.01), p=0.48 | 0.00 (−0.004, 0.004), p=0.87 | 0.001 (−0.01, 0.01), p=0.80 | 0.00 (−0.01, 0.01), p=1.00 |
| LVMI, g/m<sup>2</sup> | 0.00 (−0.01, 0.01), p=0.91 | −0.003 (−0.02, 0.01), p=0.76 | 0.01 (−0.01, 0.03), p=0.45 | 0.01 (−0.02, 0.03), p=0.56 | −0.003 (−0.02, 0.01), p=0.76 | 0.001 (−0.03, 0.03), p=0.93 |
| EDV, ml | −0.001 (−0.002, 0.001), p=0.41 | −0.002 (−0.01, 0.002), p=0.39 | 0.002 (−0.003, 0.01), p=0.33 | 0.00 (−0.01, 0.004), p=0.85 | −0.002 (−0.01, 0.002), p=0.39 | 0.002 (−0.004, 0.01), p=0.50 |
| EDVI, ml/m<sup>2</sup> | −0.001 (−0.004, 0.003), p=0.80 | −0.001 (−0.01, 0.01), p=0.78 | 0.01 (−0.01, 0.02), p=0.27 | 0.001 (−0.01, 0.01), p=0.91 | −0.001 (−0.01, 0.01), p=0.78 | 0.01 (−0.01, 0.02), p=0.33 |
| ESV, ml | −0.001 (−0.004, 0.002), p=0.57 | −0.004 (−0.01, 0.003), p=0.26 | 0.002 (−0.01, 0.01), p=0.69 | −0.002 (−0.01, 0.01), p=0.62 | −0.004 (−0.01, 0.003), p=0.26 | 0.001 (−0.01, 0.01), p=0.86 |
| ESVI, ml/m<sup>2</sup> | 0.00 (−0.01, 0.01), p=0.97 | −0.004 (−0.02, 0.01), p=0.53 | 0.004 (−0.02, 0.02), p=0.70 | −0.001 (−0.02, 0.02), p=0.89 | −0.004 (−0.02, 0.01), p=0.53 | 0.003 (−0.02, 0.02), p=0.75 |
| SV, ml | −0.001 (−0.004, 0.001), p=0.40 | −0.001 (−0.01, 0.01), p=0.70 | 0.01 (−0.003, 0.01), p=0.22 | 0.00 (−0.01, 0.01), p=0.90 | −0.001 (−0.01, 0.01), p=0.70 | 0.004 (−0.01, 0.01), p=0.38 |
| SVI, ml/m<sup>2</sup> | −0.001 (−0.01, 0.01), p=0.72 | 0.001 (−0.01, 0.02), p=0.87 | 0.01 (−0.01, 0.03), p=0.19 | 0.002 (−0.01, 0.02), p=0.77 | 0.001 (−0.01, 0.02), p=0.87 | 0.01 (−0.01, 0.03), p=0.25 |
| EF, % | −0.002 (−0.02, 0.01), p=0.70 | 0.01 (−0.02, 0.03), p=0.68 | 0.01 (−0.03, 0.04), p=0.75 | 0.001 (−0.03, 0.03), p=0.97 | 0.01 (−0.02, 0.03), p=0.68 | 0.01 (−0.03, 0.04), p=0.79 |
| Cardiac index, L/min/ m<sup>2</sup> | 0.02 (−0.18, 0.22), p=0.82 | −0.18 (−0.64, 0.28), p=0.43 | −0.38 (−0.98, 0.23), p=0.21 | 0.10 (−0.11, 0.31), p=0.33 | −0.12 (−0.62, 0.38), p=0.62 | −0.66 (−1.23, −0.10), p=0.02 |
| EDV/LVM ratio | −0.04 (−0.38, 0.31), p=0.83 | 0.14 (−0.64, 0.93), p=0.71 | 0.29 (−0.77, 1.34), p=0.58 | −0.09 (−1.00, 0.81), p=0.83 | 0.14 (−0.64, 0.93), p=0.71 | 0.83 (−0.26, 1.92), p=0.13 |
| RF, % | 0.002 (−0.01, 0.01), p=0.56 | 0.002 (−0.02, 0.02), p=0.83 | 0.001 (−0.02, 0.03), p=0.93 | 0.002 (−0.02, 0.02), p=0.79 | 0.002 (−0.02, 0.02), p=0.83 | 0.003 (−0.02, 0.03), p=0.83 |
| Barlow's disease | −0.02 (−0.22, 0.18), p=0.86 | 0.21 (−0.23, 0.64), p=0.33 | 0.22 (−0.37, 0.81), p=0.45 | 0.03 (−0.19, 0.25), p=0.79 | 0.17 (−0.31, 0.64), p=0.48 | −0.02 (−0.05, 0.003), p=0.08 |
| MAD | −0.12 (−0.53, 0.29), p=0.55 | −0.30 (−1.15, 0.55), p=0.48 | 0.01 (−1.14, 1.17), p=0.98 | −0.12 (−0.53, 0.28), p=0.54 | −0.31 (−1.17, 0.55), p=0.46 | −0.001 (−1.10, 1.10), p=1.00 |
| Bileaflet prolapse | −0.19 (−0.40, 0.02), p=0.08 | −0.35 (−0.85, 0.14), p=0.15 | 0.29 (−0.39, 0.97), p=0.39 | −0.18 (−0.40, 0.03), p=0.09 | −0.40 (−0.90, 0.09), p=0.11 | 0.38 (−0.26, 1.02), p=0.23 |
| Native T1 global, ms | 0.001 (−0.003, 0.004), p=0.73 | −0.01 (−0.01, 0.002), p=0.16 | −0.01 (−0.02, 0.004), p=0.21 | −0.01 (−0.02, 0.00), p=0.06 | −0.01 (−0.01, 0.002), p=0.16 | −0.01 (−0.02, 0.01), p=0.31 |
| Native T2 global, ms | 0.02 (−0.03, 0.08), p=0.40 | 0.06 (−0.06, 0.19), p=0.31 | −0.05 (−0.21, 0.13), p=0.59 | 0.07 (−0.07, 0.22), p=0.29 | 0.06 (−0.06, 0.19), p=0.31 | 0.04 (−0.14, 0.23), p=0.63 |
| ECV global, % | 0.02 (−0.02, 0.05), p=0.34 | 0.004 (−0.08, 0.09), p=0.92 | −0.07 (−0.18, 0.05), p=0.24 | −0.02 (−0.11, 0.08), p=0.74 | 0.004 (−0.08, 0.09), p=0.92 | −0.05 (−0.17, 0.07), p=0.42 |
| LGE, ml | −0.02 (−0.06, 0.03), p=0.40 | −0.03 (−0.13, 0.07), p=0.55 | 0.02 (−0.12, 0.15), p=0.81 | −0.04 (−0.14, 0.06), p=0.45 | −0.03 (−0.13, 0.07), p=0.55 | 0.02 (−0.11, 0.15), p=0.78 |
| LGE, % | −0.01 (−0.06, 0.04), p=0.70 | −0.01 (−0.12, 0.10), p=0.87 | 0.01 (−0.14, 0.15), p=0.91 | −0.02 (−0.13, 0.09), p=0.68 | −0.01 (−0.12, 0.10), p=0.87 | 0.01 (−0.14, 0.15), p=0.93 |

\*Adjusted for age and sex
Abbreviations: BMI – body mass index, ECV – extracellular volume, EDV – end diastolic volume, EDVI – end diastolic volume index, EF – ejection fraction, ESV – end systolic volume, ESVI – end systolic volume index, LGE – late gadolinium enhancement, LVM – left ventricular mass, LVMI – left
ventricular mass index, MAD – mitral annular disjunction, MPR – myocardial perfusion reserve, MR – mitral regurgitation, RF – mitral regurgitant fraction, SV – stroke volume, SVI – stroke volume index.

In analyses adjusted for age and sex, the associations between NYHA class and MPR remained significant (β=–0.60, 95% CI –1.17 to –0.04; p=0.04). Higher heart rate was associated with lower myocardial stress perfusion (β=–0.02, 95% CI –0.03 to –0.001; p=0.03) and lower MPR (β=–0.03, 95% CI –0.04 to –0.01; p=0.01), while higher cardiac index was also associated with lower MPR (β=–0.66, 95% CI –1.23 to –0.10; p=0.02). No significant associations were observed between RF, EDV/LVM ratio, native T1, native T2, ECV, or LGE and myocardial perfusion or MPR.

## Discussion

To our knowledge, this is the first study to use quantitative CMR perfusion mapping to assess myocardial perfusion in patients with MR due to MVP. Among prospectively enrolled patients with significant MR and matched controls without MR or MVP, patients with MR exhibited higher EDV/LVM, global native T1, and ECV, as well as a substantial prevalence of LGE. However, global myocardial perfusion at rest and during adenosine stress did not differ between groups. Within patients with MR, higher NYHA class was associated with lower MPR after adjustment for age and sex.

Chronic MR increases LV preload and imposes volume overload, leading to LV dilatation, eccentric hypertrophy, and increased total SV that initially preserve forward cardiac output (17). In this setting, myocardial oxygen demand and myocardial rest perfusion are expected to rise, which theoretically could lower MPR if hyperemic perfusion does not increase to the same extent. However, in our cohort of patients with hemodynamically significant MR referred for mitral valve surgery, global myocardial perfusion per gram of tissue was preserved, despite LV volume overload. Because patients with MR had a greater LV myocardial mass, the estimated total coronary flow was higher. Thus, preserved perfusion per gram of myocardium does not imply an unchanged total coronary flow requirement; rather, a larger myocardial mass requires a greater total blood flow to maintain similar tissue-level perfusion.

The lower MPR observed in symptomatic patients together with higher native T1, ECV, and LGE, suggests that CMD may emerge in parallel with progressive remodelling and symptom development in more advanced disease stages. This pattern suggests that compensatory mechanisms, such as microvascular adaptation, increased capillary recruitment and favourable loading conditions, can sustain adequate perfusion under chronic volume overload as long as LV function is preserved. This contrasts with non-ischemic cardiomyopathies, where impaired myocardial perfusion and reduced MPR are common and have been associated with adverse clinical outcomes, suggesting that CMD may play a more primary role in disease progression (18–22).

Our results contrast with the only prior study identified, by Akasaka et al, which reported reduced coronary flow reserve (CFR) in patients with chronic primary MR. In that invasive Doppler study, CFR reduction was mainly driven by increased baseline coronary flow velocity, while maximal hyperemic flow remained unchanged, a pattern attributed to elevated LV preload, increased LV end-diastolic pressure, and LV hypertrophy (23). However, the study by Akasaka et al. differs from the present one in several important ways. First, Akasaka et al. used invasive Doppler guidewire measurements in a single coronary artery, whereas our study employed non-invasive, fully quantitative CMR perfusion mapping, providing global myocardial perfusion assessment and measuring absolute myocardial blood flow per gram of tissue, which may better account for LV hypertrophy than flow velocity-based indices (24). Second, their patient cohort appeared to represent a more advanced disease stage, with higher RF, larger LV volumes, and greater LV mass prior to surgical referral.

Against this background, the observed reduction in MPR in symptomatic patients (NYHA class > I) with MR in our cohort may indicate that these compensatory mechanisms begin to fail with increasing disease burden and symptom severity, supporting the concept that CMD is a secondary phenomenon rather than a primary driver in MR.

In line with this, beyond myocardial perfusion, we found elevated native T1, ECV, and a higher prevalence of LGE in MR patients compared with controls, indicating interstitial fibrosis and myocardial structural remodelling. These changes in tissue characteristics could, over time, impair microvascular function (20), potentially explaining why CMD becomes apparent in more symptomatic or advanced disease. Longitudinal studies assessing changes in myocardial perfusion, myocardial remodelling, and symptoms over time, particularly following mitral valve surgery, are therefore needed to clarify the temporal relationship between microvascular dysfunction, myocardial remodelling, and symptom development in primary MR.

### Limitations

This study has limitations. The relatively small sample size may limit statistical power and generalizability, particularly for subgroup analyses. In five controls, full coverage LGE was not obtained which precludes exclusion of focal fibrosis, however there were no signs of focal fibrosis in the acquired ECV images. In three controls rest perfusion imaging was not performed and myocardial stress perfusion maps were not obtained in five patients with MR and one control due to inadequate peak stress or poor image quality.

## Conclusions

In patients with significant MR due to MVP, myocardial perfusion appears preserved despite LV volume overload. However, the lower MPR observed in symptomatic patients together with higher native T1, ECV, and LGE, suggests that CMD may emerge in parallel with progressive remodelling and symptom development in more advanced disease stages. Future longitudinal studies, particularly those assessing changes in myocardial perfusion and tissue characteristics after mitral valve intervention, are needed to clarify the temporal relationship between MR severity, LV remodelling, myocardial perfusion and clinical outcomes.

## Clinical Perspectives

### Clinical implications

In patients with MVP and significant MR, global myocardial perfusion at rest and during stress, as well as MPR, are preserved despite substantial LV volume overload. These findings indicate that CMD is unlikely to be a primary driver in compensated primary MR. However, the lower MPR in symptomatic patients (NYHA class > I), together with higher native T1, ECV, and LGE, suggests that CMD may emerge in parallel with progressive remodelling and symptom development in more advanced disease stages. Quantitative CMR perfusion mapping offers a non-invasive approach to assess myocardial perfusion and may provide additional insight into the myocardial consequences of chronic volume overload in the setting of primary MR.

### Translational Outlook

Future studies are needed to determine whether quantitative CMR perfusion mapping can detect changes in myocardial perfusion following mitral valve intervention, and how these relate to reverse remodelling and symptom burden. Integrating quantitative perfusion mapping with tissue characterization in primary MR could improve monitoring of disease progression and help refine the timing of surgery in patients with primary MR, particularly around the transition from asymptomatic to symptomatic stages.

## Data Availability

The data underlying this article will be shared on reasonable request to the corresponding author.

## Abbreviations

CMD: Coronary microvascular dysfunction
CMR: Cardiovascular magnetic resonance
CFR: Coronary flow reserve
ECV: Extracellular volume
EDV: End-diastolic volume
EF: Ejection fraction
ESV: End systolic volume
FED: Fibroelastic deficiency
LGE: Late gadolinium enhancement
LV: Left ventricular
LVM: Left ventricular mass
MAD: Mitral annular disjunction
MPR: Myocardial perfusion reserve
MR: Mitral regurgitation
MVP: Mitral valve prolapse
RF: Regurgitant fraction
SV: Stroke volume

---

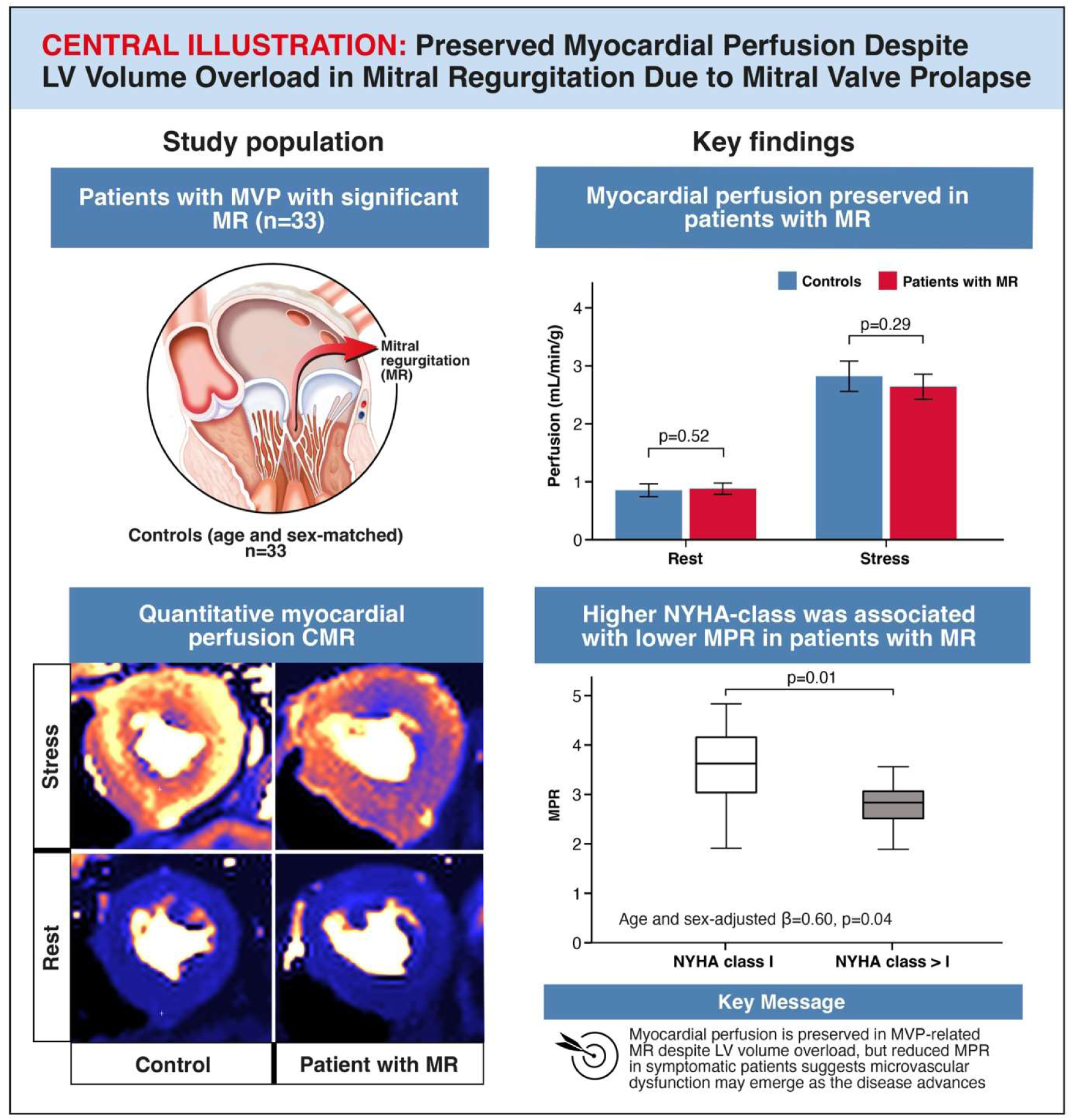
Central Illustration. Preserved Myocardial Perfusion Despite LV Volume Overload in Mitral Regurgitation Due to Mitral Valve Prolapse.

